# Transcranial Direct Current Stimulation (tDCS) Augments the Effects of Gamified, Mobile Attention Bias Modification

**DOI:** 10.1101/2020.04.20.20057141

**Authors:** Sarah Myruski, Hyein Cho, Marom Bikson, Tracy A. Dennis-Tiwary

## Abstract

Anxiety-related attentional bias (AB) is the preferential processing of threat observed in clinical and sub-clinical anxiety. Attention bias modification training (ABMT) is a computerized cognitive training technique designed to systematically direct attention away from threat and ameliorate AB, but mixed and null findings have highlighted gaps in our understanding of mechanisms underlying ABMT and how to design the most effective delivery systems. One neuromodulation technique, transcranial direct current stimulation (tDCS) across the prefrontal cortex (PFC) may augment the effects of ABMT by strengthening top-down cognitive control processes, but the evidence base is limited and has not been generalized to current approaches in digital therapeutics, such as mobile applications. The present study tested whether tDCS across the PFC, versus sham stimulation, effectively augments the beneficial effects of a gamified ABMT mobile app. Thirty-eight adults (*M*_*age*_ = 23.92, *SD* = 4.75; 18 females) evidencing low-to-moderate anxiety symptoms were randomly assigned to active or sham tDCS for 30-minutes while receiving ABMT via a mobile app. Participants reported on potential moderators of ABMT, including life stress and trait anxiety. ECG was recorded during a subsequent stressor to generate respiratory sinus arrhythmia (RSA) suppression as a metric of stress resilience. The app overall reduced subjective anxiety, whereas adding tDCS (compared to sham) reduced AB and boosted stress resilience measured via RSA suppression, particularly for those reporting low life stress. Our results integrating tDCS with ABMT provide insight into the mechanisms of AB modulation and support ongoing evaluations of enhanced ABMT reliability and effectiveness via tDCS.

---

Anxiety disorders are among the most common and costly of mental health conditions in the United States (Kessler, Berglund, Demler, Jin, Merikangas, & Walters, 2005), but only a fraction of patients seek treatment, and a third of those do not respond to current treatment options (Bystritsky, 2006). Thus, recent research and clinical efforts have focused on identifying new targets of intervention and reducing barriers to accessing treatment (e.g., Barak, Hen, Boniel-Nissim, & Shapira, 2008; Kazdin & Rabbitt, 2013; Dennis & O’Toole, 2014; Rotheram-Borus, Swendeman, & Chorpita, 2012).

A large body of evidence suggests that individuals evidencing elevated anxiety also show an attention bias (AB) for threat-relevant stimuli, or selective and exaggerated attention to and difficulty disengaging from threat-relevant stimuli (Bar-Haim, Lamy, Pergamin, Bakermans-Kranenburg, & Van Ijzendoorn, 2007; Bradley, Mogg, & Millar, 2000; Mathews & MacLeod, 2005). Techniques such as computerized attention bias modification training (ABMT) have been developed both to examine the causal nature of AB (e.g., Mathews, & MacLeod, 2002) and to serve as a computerized therapy-delivery system. Substantial evidence suggests that training anxious individuals to attend away from threat and towards non-threatening stimuli via ABMT reduces AB and anxiety severity in clinical and sub-clinical anxiety (e.g., Hakamata et al., 2010; Dennis-Tiwary et al., 2019) and across ages including children (e.g., Eldar, Ricon, & Bar-Haim, 2008; Pergamin-Hight et al., 2016), although recent meta-analyses also document small effect sizes and raise questions about clinical relevance (e.g., Cristea et al., 2015). Thus, although more evidence is needed, ABMT, which is brief, accessible, cost-effective, and low-toxicity, is a promising new anxiety- and stress-reduction intervention for people experiencing both clinical and sub-clinical anxiety, and for whom intensive treatments may be too time-consuming and cost-prohibitive.

In addition, there remain significant gaps in our understanding of mechanisms that underlie ABMT, which limits the refinement of efficacious, personalized ABMT techniques. Recent neuroimaging and neurophysiological studies point to the potential mechanistic role of prefrontal-cortex (PFC)-mediated changes in attention control (Cisler & Koster, 2010; Dennis-Tiwary et al., 2016; Heeren, De Raedt, Koster, & Philippot, 2013). Several models posit that anxiety-related AB may result from disruptions in the ability to recruit top-down attention control (e.g., Bishop, Duncan, Brett, & Lawrence, 2004; Cisler & Koster, 2010; Dennis-Tiwary, Roy, Denefrio, & Myruski, 2019; MacLeod, Mathews, & Tata, 1986), which would be signaled by reduced activation of the PFC, in particular the dorsal lateral PFC (DLPFC; Bishop, 2009; Browning, Holmes, Murphy, Goodwin, & Harmer, 2010).

Prior studies have directly tested this causal hypothesis by manipulating recruitment of the PFC during ABMT procedures using transcranial direct current stimulation (tDCS). tDCS is a portable battery-powered device that delivers low-intensity (∼2 mA) direct electric current through electrodes positioned over the scalp (Bikson et al., 2016; Woods et al., 2016). When this current reaches the neural tissue, the polarization of the resting membrane potential is shifted (Radman, Ramos, Brumberg, & Bikson, 2009) modulating ongoing plasticity (Fritsch et al., 2010; Kronberg, Bridi, Abel, Bikson, & Parra, 2017; Kronberg, Rahman, Sharma, Bikson, & Parra, 2019), for example plasticity produced by ABMT (Heeren et al., 2015). tDCS can be used both in academic and clinical centers and, when appropriate design steps are taken, at remote settings (e.g. work, home; Charvet et al., 2015).

Two recent studies provide evidence that tDCS nominally targeting the left DLPFC influences ABMT efficacy (Clarke, Browning, Hammond, Notebaert, & MacLeod, 2014; Heeren et al. 2015). For example, Clarke and colleagues (2014) selecting non-anxious participants, reported that subjects who received tDCS versus sham (placebo) while completing ABMT showed changes in attention bias in either targeted direction - both towards and away from threat. This suggests that tDCS enhances AB plasticity, however directed. A subsequent study (Heeren et al., 2015) selecting for high trait anxiety adults (who did not show AB at baseline), found that combining ABMT and tDCS reduced an eye tracking index of AB (duration of gaze fixated on threat). Taken together, these studies document that tDCS augments the impact of ABMT on AB as a target cognitive process in the etiology and maintenance of anxiety, but may interact with ABMT in an individual- and protocol-specific manner.

While computerized ABMT techniques significantly reduce treatment barriers, they are typically administered on a desktop computer in a laboratory or clinic setting, which remain difficult to access. In addition, traditional ABMT techniques are often described as repetitive and engagement and motivation by participants tends to be low (Dennis & O’Toole, 2014). To address these barriers, we have created a mobile version of ABMT that is more engaging than the traditional ABMT protocol and can be used on mobile devices. This commercially available mobile application or “app” (for iOS devices like iPhones), called Personal Zen, takes the core components of the gold-standard ABMT protocol and puts them in the context of an appealing game (see Dennis & O’Toole, 2014). It further incorporates video game-like features such as animated characters and sound effects. Like traditional ABMT, attention is still systematically redirected away from threat-relevant stimuli (angry faces).

We have recently demonstrated in two placebo-controlled studies with college students evidencing elevated trait anxiety that this user-friendly and engaging version of ABMT reduced anxiety, stress reactivity, and AB in a single, lab-based session (Dennis & O’Toole, 2014; Dennis-Tiwary et al., 2016). Moreover, in a placebo-controlled trial including pregnant women who used the app for 30-40 minutes a week for one month, stress reactivity measured via salivary cortisol and subjective anxiety were significantly reduced. These data demonstrate that the app is an effective delivery system for ABMT.

Prior ABMT studies studies (Egan & Dennis-Tiwary, 2018; Pergamin-Hight et al., 2016; Price et al., 2018) and studies utilizing Personal Zen as the delivery system for ABMT (Dennis-Tiwary et al., 2016; Dennis-Tiwary, Denefrio, & Gelber, 2017) have further documented moderators of AB plasticity, including anxiety severity and life stress (Egan & Dennis-Tiwary, 2018; Wald et al., 2016), suggesting that in order to fully understand mechanisms in ABMT, such moderators should be explored.

The present study will test whether tDCS administered during a gamified, mobile version of ABMT serves to boost beneficial effects. We will test the hypothesis that participants receiving tDCS across the PFC, compared to sham tDCS, will show augmented benefits of ABMT, measured via reduced anxious mood, reduced AB, and enhanced stress resilience [measured via respiratory sinus arrhythmia (RSA) measured during a stressor]. Individual differences in anxiety severity and life stress will be explored as potential moderators of effects.

## Methods

### Participants

Adults were screened to participate from a pool of undergraduate students and community members at an urban college campus. Participants were excluded if they reported history of epilepsy, irremovable metallic pieces in or around the head, head or neck tattoos, severe skin sensitivity or condition (e.g. eczema) affecting the face or scalp, latex allergies, history of head or traumatic brain injury, use of hearing aid devices, or pregnancy. An a priori power analysis (conducted via G*Power) showed that a sample size of 42 participants would be sufficient to detect medium effect sizes (*f* = .31 and above) across the two target within-subjects measures (pre-tDCS and post-tDCS) at 96% power. Total of 60 participants were recruited to account for potential data loss. Out of these individuals, seven participants were excluded at phone screen due to one or more of these criteria, fourteen chose not to participate following the phone screen, and one participant was excluded before consent due to presence of a fixed metal retainer in the mouth. The final sample of 38 [*M*_age_ = 23.92, *SD* = 4.75; 18 (47.4%) females] reported normal to moderate levels of anxiety according to self-report on the Depression, Anxiety, and Stress Scale (DASS-21; Henry & Crawford, 2005; *M* = 3.16, *SD* = 3.07, *Min* = 0.00, *Max* = 13.00). Thirty-four (89.5%) of participants were right-handed, 3 (7.9%) were left-handed, and 1 (2.6%) was ambidextrous. Race/ethnicity was as follows: 14 (36.8%) White, 13 (34.2%) Asian, 6 (15.8%) Black or African-American, 1 (2.6%) American Indian/Alaskan Native, 2 (5.3%) reported more than one race, and 2 (5.3%) opted to not report this information. Further, 9 (23.7%) participants were Hispanic or Latinx, 23 (60.5%) were not Hispanic or Latinx, and 6 (15.8%) opted to not report this information. Following informed consent, participants were randomly assigned to one of the two study groups: either active experimental group (*n* = 18) that receives active electrical stimulation or sham control group (*n* = 20) in which there is no stimulation. Random assignments for study groups were achieved by an online random sequence generator. Three participants were excluded from RSA analyses due to unusable recordings. The significance of findings did not change when these individuals were removed from other analyses. This study was approved by the Institutional Review Board of Hunter College, CUNY (Protocol 334490).

## Materials and Procedure

ECG was recorded throughout the entire session. Participants first completed an AB assessment (the dot probe task) followed by self-report questionnaires. Next, tDCS was applied and administered for 30 minutes which included 25 minutes of ABMT and a 5-minute break. Participants then completed an ECG baseline and AB assessment, followed by a stressful anagrams task. Finally, a third AB assessment concluded the study session. Self-reported state anxiety and mood was assessed before and after the tDCS administration and the anagrams task. In total, each study session took approximately 2.5 to 3 hours to complete.

### Transcranial Direct Current Stimulation (tDCS)

tDCS was administered using a Soterix 1×1 tDCS Limited Total Energy (LTE) Stimulator while participants completed the ABMT app. Direct current was administered via two 5 cm x 5 cm pre-saturated saline pad (SnapPad) positioned using the OLE system corresponding to (10-20 international system) F3 Anode and F4 Cathode. The OLE montages maximizes anodal-directed current to lDPFC (Seibt, Brunoni, Huang, & Bikson, 2015) while stimulating across PFC and have been suggested optimal in prior trials of ABMT (Shahbabaie, Hatami, Farhoudian, Ekhtiari, Khatibi, & Nitsche, 2018) and in trials of other anxiety-linked psychiatric disorders (Brunoni et al., 2017). The OLE approach, being automated by the head-gear (Seibt et al., 2015), also translates to home-use consistent with the broader deployable goals of our research program (Dobbs et al., 2018; Kasschau, Reisner, Sherman, Bikson, Datta, & Charvet, 2016). The intensity of tDCS was 2 mA with a 30-sec ramp up/down time. Stimulation began approximately two minutes before the initiation of ABMT to allow for acclimation to stimulation before training began. If participants reported adverse effects/discomfort that did not abate during the ramp-up period, the acclimation period was extended until discomfort alleviated, after which participants began ABMT while tDCS was simultaneously administered at 2 mA or the maximum intensity that participants indicated was tolerable. In the sham condition identical protocol was followed except stimulation current was ramped up (30-sec) and down (30-sec) at the beginning and end of the session. Because the researchers (trained personnel) were required to operate the tDCS device in order to deliver stimulation to participants properly, only participants were blinded to whether they received the active or sham tDCS.

### Mobile, Gamified Attention Bias Modification Training (ABMT)

During the neuromodulation procedure, all participants received the active version of the ABMT app, commercially available under the name Personal Zen. Participants sat comfortably at a table and were given an iPod Touch or used their iOS device (e.g., iPhone) to practice the app to ensure understanding (see Dennis-Tiwary et al., 2017). The following instructions were provided: “In this attention training app, two animated characters will appear on the screen. Shortly after, they will burrow into a hole. One of them will cause a path of grass to rustle behind it. With your finger, trace the path of the rustling grass, beginning from the burrow. Trace the grass as smoothly, quickly, and accurately as possible. At no point should you feel rushed, you should be comfortable.” Then, they were allowed to complete one practice round under the guidance of the experimenter who answered any questions about the app. For every trial, two cartoon characters (sprites), one showing an angry expression and one showing a neutral/mildly pleasant expression, appeared simultaneously on the screen for 500 ms. Next, both sprites simultaneously “burrowed” into the grass field (See Dennis-Tiwary et al., 2016; 2017 for images of the app). Then, a trail of grass appeared in the location of the non-threat character for every trial. The grass remained until participants responded by correctly tracing the grass path starting from the point at which the sprite burrowed out of sight. Participants were instructed to play the app for two 12.5 minutes sessions (∼25 min of app play total), separated by a 5-minute break. Each session consisted of approximately 40 - 45 app rounds (varied based on user speed) with 12 trials per round. Number of training trials were consistent with previously documented effective “dosages” of the app (Dennis-Tiwary et al., 2016; 2017; Dennis & O’Toole, 2014).

### The Dot Probe

The dot probe task (Bar-Haim et al., 2007; Mathews & Mackintosh, 1998) was administered immediately prior to and following administration of tDCS/ABMT procedure to measure AB. The dot probe followed parameters of the Tel-Aviv University/National Institute of Mental Health protocol. Stimuli for the dot probe task are pictures of 20 different individuals (10 males, 10 females) from the NimStim stimulus set (Tottenham et al., 2009) with one female taken from the Matsumoto and Ekman (1989) set. Stimuli were programmed using E-Prime version 2.0 (Schneider, Eschman, & Zuccolotto, 2002).

During each trial, two pictures were presented, either angry-neutral face pairs or neutral– neutral face pairs (depicting the same individual). The pictures were shown above and below a fixation cross, with 14 mm between them. The task included 120 trials [80 threat (angry faces) and neutral faces (TN) and 40 non-threat both neutral faces (NN)]. Each trial comprised: (a) 500 ms fixation, (b) 500 ms face-pair cue, which then disappears, (c) probe (target) in the former location of one of the faces until a response is made via the left or right mouse button to indicate the direction in which the arrow is pointing, and (d) 500 ms inter-trial interval. Participants were asked to respond as quickly and as accurately as possible whether the arrow was pointing to the left or the right. Probes were equally likely to appear on the top or bottom, in the location of the angry or neutral face cues and pointing to the left or the right.

### Quantifying Attention Bias (AB)

AB was measured via the dot probe task. Dot probe trials with incorrect responses were excluded from further processing and analyses. Responses faster than −2.5 *SD* from an individual’s mean and slower than +2.5 *SD* from an individual’s mean were removed. The average response time was 535 (*SD* = 101) milliseconds and the overall accuracy rate prior to training was 0.99 (*SD* = 0.01). AB was calculated in two ways. First, to quantify overall attention capture by threat, a threat bias score was computed as the average RTs for neutral probes in TN trials minus RTs for angry probes in TN trials. Second, to quantify the more specific effortful top-down inhibition of attention, a difficulty disengaging score was computed as the average RTs for neutral probes in TN trial minus RTs for neutral probes in the NN trials.

### Anagrams Task

Participants completed an anagrams task (Bishop, 2009) consisting of 40 medium to difficult mixed letter words (i.e. anagrams; e.g. RISECET = RECITES). Fourteen of the anagrams were not solvable as real words (i.e., IUTRUCE). Participants received the following instructions verbatim: “For this task you will be asked to solve 40 anagrams. You will have three minutes to complete this task. When you have solved an anagram press the spacebar to proceed. Please do this task as quickly and as accurately as possible.” Participants were also told that the solutions must be English words, and none of the solutions were proper nouns. Anagrams were presented on the computer and participants were asked to write down their solutions on a sheet of paper. After three minutes elapsed, participants received the following instructions: “Your time is up. Please hand your score sheet to the research assistant.”

### Electrocardiogram (ECG)

#### ECG Application

ECG was recorded via a Biopac MP150 wireless system (Biopac Systems, CA, USA). Three sticker-based electrodes were applied, one to each clavicle and one on the left rib and were connected via three leads to a transmitter attached to a Velcro strap which participants wore around their waists. ECG data was wirelessly transmitted to a computer to allow for ambulatory recording with Acqknowledge v4.4 software. ECG was recorded continuously during the entire study session with the exception of questionnaire completion. Manual event-markers indicated the beginning and ending of each task.

#### ECG Processing and RSA quantification

The ECG data was segmented during recording based on the onset and offset of the two baseline tasks and the anagrams tasks. Mindware 3.14 software was later used to process data, reject artifacts, and compute scores. Inter-beat intervals (IBI) were defined as the temporal distance between R-spikes, which represent the contraction of the ventricles of the heart. ECG recordings were segmented into 30-second sections, which were each manually inspected for missing or incorrectly labeled R-spikes. Segments with greater than 10% artifacts were not included in computed scores, consistent with criteria used in previous studies (e.g. Blandon, Calkins, Keane, & O’Brien, 2008). Spectral analyses used a Hamming window, and the frequency band of spontaneous respiration in adults was targeted to quantify RSA (0.120 - 0.420 Hz) consistent with prior studies (e.g. Denver, Reed, & Porges, 2007). RSA was calculated via Mindware software using the Porges (1985) method which applies an algorithm resulting in natural log transformed variance in heart rate period while accounting for respiration in units of ln(ms)^2^.

RSA suppression (ΔRSA) was quantified using residual scores, which have been used in prior studies comparing biological responses to emotional stimuli or events like a stressor (e.g. Myruski, Bonanno, Gulyayeva, Egan, & Dennis-Tiwary, 2017). Residual scores offer an advantage in comparison to subtraction scores such that residuals are more resistant to bias due to baseline inter-correlation (Weinberg, Venables, Proudfit, & Patrick, 2014). To quantify stress resilience, ΔRSA scores were computed by generating residuals with baseline RSA as the predictor and RSA in the anagrams task as the outcome. More negative ΔRSA scores indicated greater stress resilience, or greater ability to flexibly engage regulatory processes in the face of a challenge.

### Self-Report Measures

#### tDCS Sensation Scales

Participants completed the Sensation Scale (developed by the researchers) to rate their level of discomfort due to tDCS [1 = no sensation; 2 = slight sensation; 3 = tingly; 4 = slightly uncomfortable; 5 = very uncomfortable]. For all participants, this scale was completed a minimum of four times as follows: First directly after application of the tDCS apparatus, again once stimulation reached 2 mA prior to the onset of the 30-minute stimulation period, again during a break approximately half-way through the 30-minute stimulation period, and finally, after tDCS removal. For cases in which participants’ initial sensation rating was “very uncomfortable” (5), additional Sensation Scales were administered as stimulation levels were adjusted or participants habituated to the stimulation sensation. If participants opted to reduce the level of stimulation, which was only done prior to the onset of the 30-minute stimulation period, current was set to 1.5 mA and another Sensation Scale was administered after 60 seconds. If sensation rating fell below “very uncomfortable” (5) at that point, stimulation was again increased to 2 mA to aim for uniformity in voltage across participants. In these cases, an additional Sensation Scale was administered after increasing stimulation back to 2 mA. If participant rating remained at “very uncomfortable” (5), the study session was discontinued to avoid undue participant distress.

#### The Depression, Anxiety, and Stress Scale

(DASS-21; Henry & Crawford, 2005). The DASS-21 is a 21-item questionnaire that measures the severity of symptoms across three domains: depression, anxiety, and stress. Each subscale contains 7 items, scored on a 0–3 scale, and with scores ranging from 0 to 21 for each subscale. The anxiety subscale was used for the present study to evaluate the impact of individual differences anxiety symptoms on effects of tDCS combined with ABMT. A score of 4–5 indicates mild anxiety. Participants’ anxiety scores ranged from 0 to 17, with most (82%) reporting normal levels of anxiety. The DASS-21 was used to measure anxiety, depression, and stress for study recruitment.

#### State Trait Anxiety Inventory

(STAI; Spielberger, 1983). The State Trait Anxiety Inventory (STAI) is a 40-item questionnaire that assesses state (20 questions) and trait (20 questions) anxiety symptom severity. Respondents are asked to indicate the degree to which each statement reflects how they feel right now and in general using a Likert-type scale ranging from 1 (not at all) to 4 (very much so). The STAI yields total scores on two scales reflecting state and trait anxiety. The state anxiety score was used in the current study to evaluate changes in mood related to the stressor, and the trait anxiety score to assess individual differences in anxiety.

#### Recent Life Changes Questionnaire

(RLCQ; Miller & Rahe, 1997). The RLCQ consists of 91 items listing different life events experienced in the past 12 months that can cause stress and assigns a numerical value (ranging from 18 to 123) to the level or magnitude of stress the event typically causes (e.g., a vacation receives a score of 24, whereas the death of a spouse is scored 119). Scores for every item endorsed were summed and used in analyses below to examine the impact of life stress on effects of tDCS combined with ABMT.

#### Analog Mood Scale

(AMS; MacLeod, Rutherford, Campbell, Ebsworthy, & Holder, 2002). The AMS is a brief measure of positive and negative mood consisting of three questions (i.e. “How anxious are you?”, “How sad are you?”, and “How happy are you?”). Participants were asked to indicate their present mood by identifying a location on a horizontal line divided into 30 equally sized sections labeled 1 (not at all) to 30 (very much). The AMS anxiety question was used in the current study to quantify changes in mood induced by tDCS administration and the anagrams task. Ratings were collected at baseline, following tDCS/ABMT administration, immediately prior to the stressor, and immediately following the stressor. An average of the two anxiety self-ratings prior to the stressor was calculated to examine effects of ABMT and tDCS on anxious mood.

#### tDCS Adverse Effects Questionnaire

At the conclusion of the study session, participants completed the tDCS Adverse Effects Questionnaire (Brunoni, Amadera, Berbel, Volz, Rizzerio, & Fregni, 2011) regarding their current physical, cognitive, and emotional state. Participants reported the severity of ten adverse effects such as headache, burning sensation, trouble concentrating, and acute mood change on a scale from 1 (absent) to 4 (severe). If an adverse effect was present [rating of 2 (mild) and above], participants reported the degree of relatedness the effect had to tDCS stimulation on a scale from (1) none to (5) definite.

## Results

### Descriptive statistics

Table 1 presents descriptive statistics for study variables, separately for each tDCS group (active, sham). No baseline group differences reached significance, and tDCS groups were similar in age and gender. Further, there were no significant differences between the active and sham groups regarding in adverse events (*p’s* > .05).

**Table 1.** Descriptive Statistics on Self-reported questionnaires, Attentional bias, Disengagement, and RSA

|  | Active (*n* = 18) |  | Sham (*n* = 20) |  |
| --- | --- | --- | --- | --- |
|  | Females (*n* = 9) | Males (*n* = 9) | Females (*n* = 9) | Males (*n* = 11) |
|  | *M* (*SD*) | *M* (*SD*) | *M* (*SD*) | *M* (*SD*) |
| *Age* | 23.33 (4.95) | 23.33 (4.77) | 24.22 (4.18) | 24.64 (5.52) |
| *DASS 21* |  |  |  |  |
| Depression | 6.78 (4.12) | 4.67 (6.42) | 3.11 (2.98) | 5.81 (4.17) |
| Anxiety | 4.22 (2.91) | 2.67 (4.06) | 2.44 (2.46) | 3.27 (2.90) |
| Stress | 7.44 (5.10) | 4.89 (4.88) | 4.11 (4.23) | 8.00 (3.61) |
| *STAI* |  |  |  |  |
| Trait | 48.67 (13.72) | 37.67 (12.25) | 38.22 (8.87) | 44.55 (8.35) |
| State (pre-tDCS) | 31.88 (11.05) | 28.78 (5.59) | 34.11 (12.75) | 37.00 (10.21) |
| State (post-tDCS) | 31.50 (8.16) | 31.22 (8.88) | 30.78 (9.55) | 35.00 (6.84) |
| *RLCQ* | 434.44 (137.12) | 441.67 (244.03) | 454.00 (255.53) | 439.36 (248.15) |
| *AMS* |  |  |  |  |
| Anxiety (pre-tDCS) | 7.67 (6.00) | 4.78 (3.073) | 9.33 (8.35) | 13.64 (8.82) |
| Anxiety (post-tDCS) | 3.88 (2.42) | 3.38 (3.020) | 4.33 (4.55) | 8.86 (7.52) |
| *Sensation scales* | 3.15 (0.64) | 2.32 (0.49) | 2.54 (0.52) | 2.77 (0.92) |
| *tDCS Adverse Effects* |  |  |  |  |
| Severity | 1.43(0.42) | 1.33(0.45) | 1.18(0.14) | 1.20(0.19) |
| Relationship | 1.62(0.75) | 1.25(0.38) | 1.52(0.58) | 1.27(0.32) |
| *Attentional bias* |  |  |  |  |
| Baseline (pre-tDCS) | 8.11 (23.40) | .11 (27.79) | -2.22 (20.64) | -9.36 (21.20) |
| Pre-stressor (post-tDCS) | -4.38 (11.60) | 4.33 (13.067) | -1.13 (18.92) | 8.64 (24.70) |
| Post-stressor (post-tDCS) | -1.88 (15.38) | 4.38 (16.34) | 1.13 (12.94) | 3.73 (15.52) |
| *Disengagement* |  |  |  |  |
| Baseline (pre-tDCS) | 6.67 (26.08) | 9.44 (20.60) | -3.56 (12.43) | 1.55 (28.14) |
| Pre-stressor (post-tDCS) | -12.88 (12.96) | -12.44 (28.47) | -2.00 (11.89) | 7.64 (27.90) |
| Post-stressor (post-tDCS) | 1.75 (16.08) | 13.25 (13.96) | -3.38 (31.65) | -1.64 (15.77) |
| *RSA* | 6.92 (.55) | 6.96 (.80) | 6.01 (1.40) | 6.69 (1.19) |

Note. Abbreviations are as follows: DASS-21(Depression Anxiety and Stress Scales), STAI (State-Trait Anxiety Inventory), RLCQ (Recent Life Changes Questionnaire), AMS (Analog Mood Scale), and RSA (Respiratory sinus arrhythmia).

Table 2 shows correlations among study variables (DASS-Anxiety, dot probe attention bias, dot probe disengagement, state anxiety, RSA at baseline, and baseline AMS). Measures of anxiety were significantly positively inter-correlated. Threat bias was not significantly correlated with any measures of anxiety, but RSA at baseline was significantly negatively correlated with post-stressor state anxiety.

**Table 2.** Correlations between scores of self-reported questionnaires, Attentional Bias (AB), Disengagement (DIS), and Respiratory Sinus Arrhythmia (RSA) separated by time points

|  | 1 | 2 | 3 | 4 | 5 | 6 | 7 | 8 | 9 | 10 | 11 | 12 |
| --- | --- | --- | --- | --- | --- | --- | --- | --- | --- | --- | --- | --- |
| 1.DASS-A | - |  |  |  |  |  |  |  |  |  |  |  |
| *Baseline* |  |  |  |  |  |  |  |  |  |  |  |  |
| 2.AMS-A | .22 | - |  |  |  |  |  |  |  |  |  |  |
| 3.STAI-S | .34* | .83** | - |  |  |  |  |  |  |  |  |  |
| 4.AB | -.11 | -.204 | .16 | - |  |  |  |  |  |  |  |  |
| 5.DIS | -.11 | -.38* | -.075 | .59** | - |  |  |  |  |  |  |  |
| 6.RSA | -.20 | -.048 | -.15 | -.015 | .080 | - |  |  |  |  |  |  |
| *Pre-Stressor* |  |  |  |  |  |  |  |  |  |  |  |  |
| 7. STAI-S | .69** | .43** | .53 | -.084 | -.043 | -.12 | - |  |  |  |  |  |
| 8. AB | -.053 | .077 | .049 | .070 | .27 | .16 | -.01 | - |  |  |  |  |
| 9. DIS | .33 | .054 | .030 | -.088 | .11 | -.19 | .34* | .46** | - |  |  |  |
| *Post-Stressor* |  |  |  |  |  |  |  |  |  |  |  |  |
| 10.STAI-S | .65** | .35* | .52** | .059 | .004 | -.44** | .72** | -.13 | .28 | - |  |  |
| 11.AB | .17 | -.058 | -.013 | -.21 | .14 | -.094 | .10 | .069 | .23 | -.061 | - |  |
| 12.DIS | .11 | -.080 | -.081 | -.28 | .035 | -.018 | -.027 | -.035 | .058 | -.12 | .58** | - |

Note. A: Anxiety, S: State; \* $p < .05$ . \*\* $p < .01$

**Table 3.** Regression Model – Effects of Group and Life Stress on RSA Suppression

| | *b* | *SE B* | *B* | *R*<sup>2</sup> | $\Delta R^2$ | *p* |
| --- | --- | --- | --- | --- | --- | --- |
| Model |  |  |  | .44 |  | .086 |
| Step 1 |  |  |  | .004 | .004 |  |
| Group | .13 | .55 | .064 |  |  | .71 |
| Step 2 |  |  |  | .006 | .002 |  |
| Group | .13 | .34 | .065 |  |  | .72 |
| RLS | .00 | .001 | -.046 |  |  | .80 |
| Step 3 |  |  |  | .19 | .18 |  |
| Group | 1.86 | .73 | .95 |  |  | .016 |
| RLS | .01 | .003 | 1.43 |  |  | .020 |
| Group x RLS | -.004 | .001 | -1.78 |  |  | .013 |

Note. Recent Life Stressor = RLS.

### Manipulation check: Stressor effects

To confirm that the anagrams task induced subjective anxiety, we conducted a simple within-subjects paired *t*-test between state anxiety (STAI-state) assessed immediately following the stressor (*M* = 37.54, *SD* = 11.85) compared to state anxiety immediately prior to the stressor [(*M* = 32.95, *SD* = 10.03)], *t* (36) = −3.35, *p* =.002] and at baseline [(*M* = 33.33, *SD* = 10.39)], *t* (36) = −2.36, *p* = .024]. Both *t*-tests reached significance while controlling for multiple comparisons (Bonferroni’s adjusted *p* = .025), confirming the induction of anxiety.

### Manipulation check: ABMT effects independent of tDCS

To test whether ABMT, independent of tDCS, reduced two key targets of ABMT – AB (threat bias and difficulty disengaging) and subjective anxiety (AMS-anxiety) - we conducted two 2(Time: pre, post) x 2(Sex: male, female) repeated measures ANOVAs separately for each of the two DVs.Bonferroni’s correction was applied to control for multiple comparisons for follow-up tests.

There was a significant effect of Time on AMS-anxiety, *F*(1, 34) = 9.95, *p* = .003, □ _*p*_^2^ =.226, showing that using the app reduced subjective anxiety (pre: *M* = 9.11, *SD* = 7.56; post: *M*= 5.40, *SD* = 5.43).

No significant effects emerged for AB metrics (*p’s* > .10).

### Effects of tDCS combined with ABMT

To test the main study hypothesis that participants receiving tDCS across the PFC, compared to sham tDCS, will show augmented benefits of ABMT, measured via reduced AB, reduced anxious mood (AMS-anxiety), and enhanced stress resilience (greater RSA suppression), we conducted two 2 (tDCS Group: active versus sham) x 2 (Sex: male, female) ANCOVAs with pre-tDCS measures of each DV as the covariate, separately for each AB score (threat bias and difficulty disengaging) and AMS-anxiety. Because RSA suppression is calculated as a differences score using residuals scores involving baseline RSA, baseline RSA was not used as a covariate, and an ANOVA with tDCS Group and Sex as the between-subjects factors was conducted instead for RSA as the dependent variable. Bonferroni’s correction was applied to control for multiple comparisons for follow-up tests.

The main effect of Group, *F*(1,31) = 5.18, *p* = .030, □_*p*_^2^ = .143, showed that AB,measured as difficulty disengaging, following ABMT was significantly lower in the active versus sham Group, (active: *M* = −12.65, *SD* = 21.88; sham: *M* = 3.58, *SD* = 22.62; *p* = .030). There were no significant effects on other metrics of AB (*p’s >* .10), and analyses with subjective anxiety and RSA suppression as the dependent variables did not reach significance (*p’s >* .10).

### Exploratory analyses of moderators of efficacy

To further examine individual differences in the impact of tDCS combined with ABMT on target outcomes, we conducted six hierarchical linear regressions via the SPSS PROCESS macro to test for the moderating effect of trait anxiety (STAI trait) and life stress (recent life changes over the past year) on the DVs (AMS-anxiety, AB, and RSA suppression). Step 1 was Group (active, sham); Step 2 was the moderator (recent life stressor or trait anxiety scores); and Step 3 was the interaction between the two. For AMS-anxiety and AB, pre-stressor baseline measures were included in the model as covariates. To account for multiple comparisons, the Benjamini-Hochberg procedure (Benjamini & Hochberg, 1995) was applied. All *p*-values reported below are raw and were significant using a false discovery rate of 0.10.

Group significantly predicted RSA suppression such that those in the active group showed significantly greater RSA suppression [β = .95, *p =* .016] overall. Lower levels of recent stressful life events also significantly predicted greater RSA suppression [β =1.43, *p* = .020] for the sample as a whole. Finally, there was a significant interaction [*R*^*2*^_change_ = .18, *p* = .013] such that active tDCS was associated with increased RSA suppression for low levels of recent stressful life events [β = .92, *t*(31) = 2.11, *p* = .043, Figure 1].

**Figure 1.**
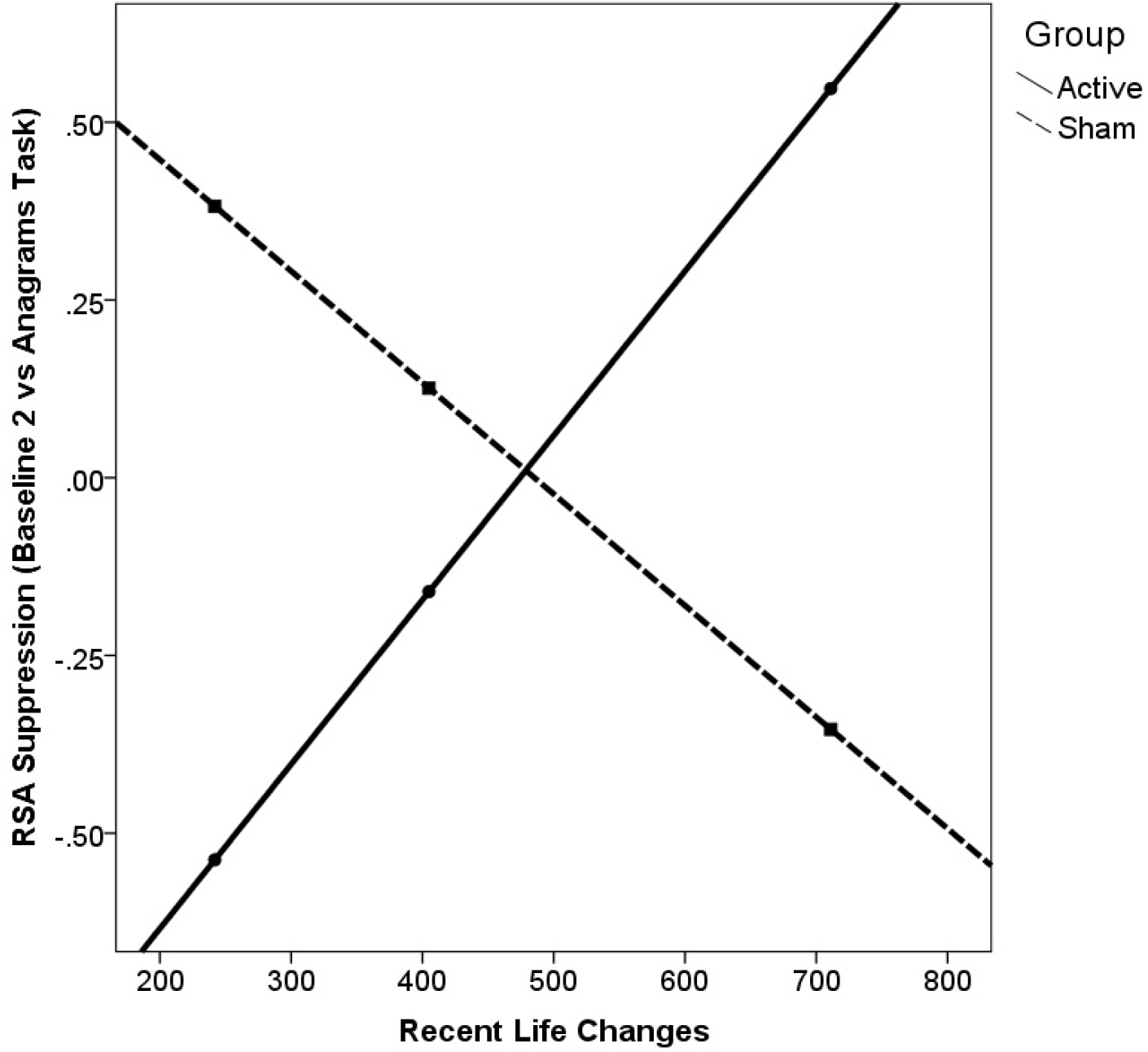
Active tDCS was associated with greater RSA suppression, but only for those with low levels of recent life changes.

No other regression analyses reached significance (model *p’s* > .10).

### Exploratory analyses of AB mediation of anxiety

We also conducted mediation analyses via the SPSS PROCESS macro to test whether AB (threat bias and difficulty disengaging) mediated the association between tDCS Group (active versus sham) and outcomes (AMS-anxiety and RSA suppression). No models reached significance (model *p’s* > .10).

## Discussion

The current study tested whether tDCS across the PFC augmented the efficacy of mobile, gamified ABMT for anxiety-related AB. It extends previous studies by using an alternative delivery system for AMBT, a mobile, gamified app, and by identifying underlying mechanisms that drive individual difference in response to ABMT. Consistent with our predictions, ABMT alone reduced subjective anxiety, whereas adding tDCS (compared to sham) reduced AB measured as difficulty disengaging attention from threat and boosted stress resilience measured via RSA suppression, particularly among those reporting lower life stress. Results provide important experimental evidence for potential mechanisms of ABMT, and advance our understanding and identification of treatment moderators, both of which support the aim of developing more personalized treatment approaches for anxiety.

Following early enthusiasm for ABMT, due to robust effect sizes emerging from well-controlled randomized clinical trials (Hakamata et al., 2010), subsequent RCTs showed mixed and null findings (Kruijt, Parsons, & Fox, 2019), leading to significant debate about the clinical utility of AB (Emmelkamp, 2012) and the potential heterogeneity of both AB and of ABMT response (Dennis-Tiwary et al., 2019). Our study documented that targeted stimulation of the PFC in combination with ABMT specifically reduced difficulty disengaging from threat, an aspect of AB closely linked to inhibitory cognitive control processes, but not threat bias, which is more closely linked to attention capture by threat (Cisler & Koster, 2010). This differential finding strengthens the evidence base that changes in PFC-mediated cognitive control processes may underlie the positive effects of ABMT, particularly on AB.

Current findings are also in line with previous data suggesting that PFC activation may modulate difficulty disengaging attention from threat among high-anxious individuals. For instance, highly trait-anxious individuals reporting poor attention control (as a proxy of the reduced PFC activity) exhibit more delayed disengagement from threat (Derryberry & Reed, 2002). Consistently, at the neural level, cortical structures centered around the prefrontal cortex and its functionally related structures (i.e., anterior cingulate cortex and orbitofrontal cortex) may mediate delayed disengagement from threat through individual differences in the ability to down-regulate the influence of limbic structures and maintain attention on task-relevant stimuli (Bishop, 2009; Blair et al., 2012). This hypothesis makes sense in the context of previous work demonstrating that the activation of the PFC is functionally related to a down-regulation of amygdala activity during the presentation of threatening stimuli (Bishop et al., 2004). Future studies should examine the impact of tDCS across the PFC during ABMT using neuroimaging techniques such as fMRI to explore whether such activation is associated with modulation of PFC—amygdala connectivity during the dot-probe task or of specific regions of the cortex, such as the lateral PFC (Browning et al., 2010).

As predicted, we also found that tDCS combined with ABMT led to greater RSA suppression in response to a stressor. Larger magnitude RSA suppression is considered an adaptive physiological response reflecting the ability to flexibly shift between parasympathetic and sympathetic nervous system engagement in the service of coping with an emotional challenge. This finding is novel and suggests that downstream indices of affective regulation may be impacted directly by tDCS. In particular, bifrontal tDCS may target cortical pathways responsible for descending control of the autonomic nervous system, which include the insular cortex and anterior cingulate cortex as identified in human imaging studies (see Cechetto, 2014, for a review).

Importantly, these effects were particularly robust among those reporting low levels of current life stress. In contrast, active tDCS did not predict greater RSA suppression among those with relatively high recent life stress, potentially due to the deleterious effect of life stressors both severe (e.g. Valerio, 2004) and moderate (Jarczok et al., 2013) on ANS activity which may have overshadowed any bolstering effects of tDCS. This finding points to the need for more research exploring the impact of treatment moderators, both general and specific to either ABMT or tDCS. Such an evidence base is a necessary first step towards truly personalized treatment approaches and clinical specificity in identifying those who are most likely to benefit from both treatment approaches (Dennis-Tiwary et al., 2019; Heeren et al., 2015; Price et al. 2018).

This study was unique in its use of mobile, gamified ABMT combined with tDCS, which cannot be directly compared with prior studies combining tDCS and ABMT (Clarke et al., 2014; Heeren et al., 2015). However, the evidence base for this particular app is promising (Dennis & O’Toole, 2014; Dennis-Tiwary et al., 2016; 2017; Sprunger, 2018), and with the growth of evidence-based digital therapeutics that reduce barriers to treatment access (Kazdin & Blase, 2011; Kazdin & Rabbitt, 2013), it remains a key goal to empirically test the generalizability of digital mental health interventions, including mobile applications.

In addition to the relatively small sample size, and challenges in the reliable assessment of AB (e.g., Waechter & Stolz, 2015), several limitations should be noted. First, while tDCS was compared to sham stimulation, ABMT was not compared to a placebo control. While this limited our ability to examine interactions between ABMT and tDCS, we were able to demonstrate that across both tDCS conditions, subjective anxiety was reduced after using the app, consistent with previous studies (Dennis & O’Toole, 2014; Dennis-Tiwary et al., 2016; 2017). This suggests that regardless of neurocognitive changes induced by tDCS, ABMT may reduce subjective negative mood specifically, but future research with a placebo control is needed. Moreover, we did not investigate the effects of tDCS without ABMT, and thus cannot rule out the possibility that tDCS alone, without ABMT, reduces AB and promotes RSA suppression. Relevant to this question, however, a prior study (Coussement et al., 2019) tested whether tDCS alone induced plasticity in AB, reasoning that strengthening cognitive control via tDCS might directly reduce AB, which is thought to be characterized by inadequate cognitive control. This study reported null findings, suggesting that tDCS alone is not sufficient to modify AB. On the other hand, participants in this study, much like the present study and a prior study combining tDCS and ABMT (Clarke et al., 2014), did not exhibit elevated AB at baseline, which may indicate that the presence of significant AB is a necessary condition for AB plasticity especially in the absence of interventions intended to train AB, such as ABMT (e.g., Heeren et al., 2015; Mogoaşe, David, & Koster, 2014). Indeed, findings of Clarke and colleagues (2014) showed an effect of tDCS on training to avoid threat and to attend to threat (the latter at the level of a trend), indicating that tDCS may more broadly influence the modification of both AB towards and away from threat. Thus, results should be interpreted with caution and future research could benefit from including both control conditions simultaneously for ABMT and tDCS, to directly cross both the presence/absence of ABMT with tDCS, and to examine the effects of each alone.

We also did not compare distinct tDCS montages, as has been done in prior research which compared anodal stimulation across the left versus right dlPFC (Heeren et al., 2015), or examine alternative position of the reference electrode (cathode), which may influence the current flow pattern through the brain (Bikson, Datta, Rahman, & Scaturro, 2010). While high-definition tDCS (HD-tDCS; Datta, Bansal, Diaz, Patel, Reato, & Bikson, 2009) allows targeting of specific cortical regions such as lDLPFC (Hill, Rogasch, Fitzgerald, & Hoy, 2018; Martínez-Pérez, Castillo, Sánchez-Pérez, Vivas, Campoy, & Fuentes, 2019; Nikolin, Loo, Bai, Dokos, & Martin, 2015; Shen, Yin, Wang, Zhou, McClure, & Li, 2016), we adopted an approach activating PFC which is broadly implicated in ABMT, and also to support future home-use of tDCS (Shaw et al., 2017) in combination with mobile ABMT.

Because tDCS is thought to boost learning (Buch et al., 2017; Kronberg et al., 2019; O’Shea et al., 2014), it is important to distinguish between effects of tDCS on acute states (such as stress) and effects related to treatment such as ABMT. That is, if may be that tDCS boosts response only if an individual is already primed to benefit from a particular treatment (Bikson & Rahman, 2013), making tDCS an enhancer of responsiveness. A complementary question about the enhancement perspective on tDCS is whether moderators – such as anxiety and life stress explored in the current study – serve as general predictors of who responds to ABMT, and thus will also predict who responds best to tDCS combined with ABMT. On the other hand, it is unknown whether some moderators specifically that predict relative benefit of tDCS versus sham, rather than overall responsiveness. Identification of the latter would represent a key inclusion criterion in a pivotal trial of tDCS versus sham.

Another limitation was that participants evidenced on average low to moderate levels of anxiety and received only a single session of ABMT. Both of these factors may have reduced our ability to detect additional effects of ABMT, which has shown mixed efficacy across studies with different populations and methods (e.g., Cristea et al., 2015) and may be most effective when administered over multiple, weekly sessions for clinically relevant elevations in AB and anxiety (e.g., Hakamata et al., 2010). Lower levels of anxiety severity, along with the relatively small sample size, might also have limited our ability to detect whether changes in AB mediated the effects of tDCS on target outcomes. Moreover, while AB is evidenced in both clinical and non-clinical anxiety (Bar-Haim et al., 2007), much of the compelling evidence for ABMT efficacy includes patients diagnosed with social anxiety or generalized anxiety disorders (Mogoaşe et al., 2014; Hakamata et al., 2010). Thus, it remains unclear whether current study findings can generalize to clinical populations. Moreover, the relatively low-to-moderate levels of anxiety severity in the current sample may have also limited our ability to detect moderating effects of anxiety on the efficacy of ABMT and/or tDCS. At the same time, by documenting the potential for tDCS to augment effects of ABMT even among those with relatively few anxiety symptoms, current findings suggests that ABMT can be enhanced to be broadly effective across the full spectrum of anxiety. Future research should include a broader range of anxiety severity including the clinical range, and explore additional moderators of efficacy.

Taken together, findings document that tDCS across the PFC combined with mobile ABMT reduced AB measured as difficulty disengaging attention from threat and boosted stress resilience measured via RSA suppression, particularly among those reporting lower life stress. Results lay the groundwork for crucial assessments of dose response parameters and more targeted examination of treatment mechanisms, as well as the identification of treatment moderators and the development of more personalized treatment approaches.

## Data Availability

The data that support the findings of this study are available from the corresponding author upon reasonable request.

## Acknowledgments

This research was made possible by the following funding sources granted to Tracy Dennis-Tiwary: National Institute of Mental Health (R56MH111700, RF1MH120846) and the National Center for Advancing Translational Sciences of the National Institutes of Health (TR000457). This research was also made possible by funding sources granted to Marom Bikson: National Institutes of Health (NIH-NIMH 1R01MH111896, NIH-NIMH 1R01MH109289; NIH-NINDS 1R01NS101362).

## References

Bar-Haim, Y., Lamy, D., Pergamin, L., Bakermans-Kranenburg, M. J., & Van Ijzendoorn, M. H. (2007). Threat-related attentional bias in anxious and nonanxious individuals: a meta-analytic study. Psychological Bulletin, 133(1), 1–24.

Barak, A., Hen, L., Boniel-Nissim, M., & Shapira, N. A. (2008). A comprehensive review and a meta-analysis of the effectiveness of internet-based psychotherapeutic interventions. Journal of Technology in Human services, 26(2-4), 109–160.

Benjamini, Y., & Hochberg, Y. (1995). Controlling the false discovery rate: a practical and powerful approach to multiple testing. Journal of the Royal statistical society: series B (Methodological), 57(1), 289–300.

Bradley, B. P., Mogg, K., & Millar, N. H. (2000). Covert and overt orienting of attention to emotional faces in anxiety. Cognition & Emotion, 14(6), 789–808.

Bikson, M., Datta, A., Rahman, A., & Scaturro, J. (2010). Electrode montages for tDCS and weak transcranial electrical stimulation: role of “return” electrode’s position and size. Clinical neurophysiology: official journal of the International Federation of Clinical Neurophysiology, 121(12), 1976.

Bikson, M., Grossman, P., Thomas, C., Zannou, A. L., Jiang, J., Adnan, T., … & Brunoni, A. R. (2016). Safety of transcranial direct current stimulation: evidence based update 2016. Brain Stimulation, 9(5), 641–661.

Bikson, M., & Rahman, A. (2013). Origins of specificity during tDCS: anatomical, activity-selective, and input-bias mechanisms. Frontiers in Human Neuroscience, 7, 688.

Bishop, S., Duncan, J., Brett, M., & Lawrence, A. D. (2004). Prefrontal cortical function and anxiety: controlling attention to threat-related stimuli. Nature Neuroscience, 7(2), 184.

Bishop, S. J. (2009). Trait anxiety and impoverished prefrontal control of attention. Nature Neuroscience, 12(1), 92.

Blair, K. S., Geraci, M., Smith, B. W., Hollon, N., DeVido, J., Otero, M., … & Pine, D. S. (2012). Reduced dorsal anterior cingulate cortical activity during emotional regulation and top-down attentional control in generalized social phobia, generalized anxiety disorder, and comorbid generalized social phobia/generalized anxiety disorder. Biological Psychiatry, 72(6), 476–482.

Blandon, A. Y., Calkins, S. D., Keane, S. P., & O’Brien, M. (2008). Individual differences in trajectories of emotion regulation processes: The effects of maternal depressive symptomatology and children’s physiological regulation. Developmental Psychology, 44(4), 1110–1131.

Browning, M., Holmes, E. A., Murphy, S. E., Goodwin, G. M., & Harmer, C. J. (2010). Lateral prefrontal cortex mediates the cognitive modification of attentional bias. Biological psychiatry, 67(10), 919–925.

Brunoni, A. R., Amadera, J., Berbel, B., Volz, M. S., Rizzerio, B. G., & Fregni, F. (2011). A systematic review on reporting and assessment of adverse effects associated with transcranial direct current stimulation. International Journal of Neuropsychopharmacology, 14(8), 1133–1145.

Brunoni, A. R., Moffa, A. H., Sampaio-Junior, B., Borrione, L., Moreno, M. L., Fernandes, R. A., … & Chamorro, R. (2017). Trial of electrical direct-current therapy versus escitalopram for depression. New England Journal of Medicine, 376(26), 2523–2533.

Buch, E. R., Santarnecchi, E., Antal, A., Born, J., Celnik, P. A., Classen, J., … & Pascual-Leone, A. (2017). Effects of tDCS on motor learning and memory formation: a consensus and critical position paper. Clinical Neurophysiology, 128(4), 589–603.

Bystritsky, A. (2006). Treatment-resistant anxiety disorders. Molecular Psychiatry, 11(9), 805. Cechetto, D. F. (2014). Cortical control of the autonomic nervous system. Experimental Physiology, 99(2), 326–331.

Charvet, L. E., Kasschau, M., Datta, A., Knotkova, H., Stevens, M. C., Alonzo, A., … & Bikson, M. (2015). Remotely-supervised transcranial direct current stimulation (tDCS) for clinical trials: guidelines for technology and protocols. Frontiers in Systems Neuroscience, 9, 26.

Cisler, J. M., & Koster, E. H. (2010). Mechanisms of attentional biases towards threat in anxiety disorders: An integrative review. Clinical Psychology Review, 30(2), 203–216.

Clarke, P. J., Browning, M., Hammond, G., Notebaert, L., & MacLeod, C. (2014). The causal role of the dorsolateral prefrontal cortex in the modification of attentional bias: evidence from transcranial direct current stimulation. Biological Psychiatry, 76(12), 946–952.

Coussement, C., Maurage, P., Billieux, J., & Heeren, A. (2019). Does change in attention control mediate the impact of tDCS on attentional bias for threat? Limited evidence from a double-blind sham-controlled experiment in an unselected sample. Psychologica Belgica, 59(1).

Cristea, I.A., Kok, R.N., & Cuijpers (2015). Efficacy of cognitive bias modification interventions in anxiety and depression: meta-analysis. British Journal of Psychiatry, 206 (1), 7–16.

Datta, A., Bansal, V., Diaz, J., Patel, J., Reato, D., & Bikson, M. (2009). Gyri-precise head model of transcranial direct current stimulation: improved spatial focality using a ring electrode versus conventional rectangular pad. Brain Stimulation, 2(4), 201–207.

Derryberry, D., & Reed, M. A. (2002). Anxiety-related attentional biases and their regulation by attentional control. Journal of Abnormal Psychology, 111(2), 225.

Dennis-Tiwary, T., Egan, L.J., Babkirk, S., and Denefrio, S. (2016). For whom the bell tolls: Neurocognitive individual differences in the acute stress-reduction effects of an attention bias modification game for anxiety. Behaviour Research and Therapy, 77, 105–117.

Dennis, T.A., & O’Toole, L. (2014). Mental health on the go: Effects of a gamified attention bias modification mobile application in trait anxious adults. Clinical Psychological Science, 2(2), 1–15.

Dennis-Tiwary, T. A., Roy, A. K., Denefrio, S., & Myruski, S. (2019). Heterogeneity of the Anxiety-Related Attention Bias: A Review and Working Model for Future Research. Clinical Psychological Science, 7(5), 879–899.

Denver, J. W., Reed, S. F., & Porges, S. W. (2007). Methodological issues in the quantification of respiratory sinus arrhythmia. Biological Psychology, 74(2), 286–294.

Dobbs, B., Pawlak, N., Biagioni, M., Agarwal, S., Shaw, M., Pilloni, G., … & Charvet, L. (2018). Generalizing remotely supervised transcranial direct current stimulation (tDCS): feasibility and benefit in Parkinson’s disease. Journal of Neuroengineering and Rehabilitation, 15(1), 114.

Fritsch, B., Reis, J., Martinowich, K., Schambra, H. M., Ji, Y., Cohen, L. G., & Lu, B. (2010). Direct current stimulation promotes BDNF-dependent synaptic plasticity: potential implications for motor learning. Neuron, 66(2), 198–204.

Egan, L. J., & Dennis-Tiwary, T. A. (2018). Dynamic measures of anxiety-related threat bias: Links to stress reactivity. Motivation and Emotion, 42(4), 546–554.

Eldar, S., Ricon, T., & Bar-Haim, Y. (2008). Plasticity in attention: Implications for stress response in children. Behaviour Research and Therapy, 46(4), 450–461.

Emmelkamp, P. M. (2012). Attention bias modification: the Emperor’s new suit?. BMC Medicine, 10(1), 63.

Hakamata, Y., Lissek, S., Bar-Haim, Y., Britton, J. C., Fox, N. A., Leibenluft, E., … & Pine, D.S. (2010). Attention bias modification treatment: a meta-analysis toward the establishment of novel treatment for anxiety. Biological Psychiatry, 68(11), 982–990.

Heeren, A., Baeken, C., Vanderhasselt, M. A., Philippot, P., & De Raedt, R. (2015). Impact of anodal and cathodal transcranial direct current stimulation over the left dorsolateral prefrontal cortex during attention bias modification: an eye-tracking study. PLoSOne, 10(4), e0124182.

Heeren, A., De Raedt, R., Koster, E. H., & Philippot, P. (2013). The (neuro) cognitive mechanisms behind attention bias modification in anxiety: proposals based on theoretical accounts of attentional bias. Frontiers in Human Neuroscience, 7, 119.

Hill, A. T., Rogasch, N. C., Fitzgerald, P. B., & Hoy, K. E. (2018). Effects of single versus dual-site High-Definition transcranial direct current stimulation (HD-tDCS) on cortical reactivity and working memory performance in healthy subjects. Brain Stimulation, 11(5), 1033–1043.

Henry, J. D., & Crawford, J. R. (2005). The short-form version of the Depression Anxiety Stress Scales (DASS-21): Construct validity and normative data in a large non-clinical sample. British Journal of Clinical Psychology, 44(2), 227–239.

Jarczok, M. N., Jarczok, M., Mauss, D., Koenig, J., Li, J., Herr, R. M., & Thayer, J. F. (2013). Autonomic nervous system activity and workplace stressors—a systematic review. Neuroscience & Biobehavioral Reviews, 37(8), 1810–1823.

Kasschau, M., Reisner, J., Sherman, K., Bikson, M., Datta, A., & Charvet, L. E. (2016). Transcranial direct current stimulation is feasible for remotely supervised home delivery in multiple sclerosis. Neuromodulation: Technology at the Neural Interface, 19(8), 824–831.

Kazdin, A. E., & Blase, S. L. (2011). Interventions and models of their delivery to reduce the burden of mental illness: Reply to commentaries. Perspectives on Psychological Science, 6(5), 507–510.

Kazdin, A. E., & Rabbitt, S. M. (2013). Novel models for delivering mental health services and reducing the burdens of mental illness. Clinical Psychological Science, 1(2), 170–191.

Kessler, R. C., Berglund, P., Demler, O., Jin, R., Merikangas, K. R., & Walters, E. E. (2005). Lifetime prevalence and age-of-onset distributions of DSM-IV disorders in the National Comorbidity Survey Replication. Archives of General Psychiatry, 62(6), 593–602.

Kronberg, G., Bridi, M., Abel, T., Bikson, M., & Parra, L. C. (2017). Direct current stimulation modulates LTP and LTD: activity dependence and dendritic effects. Brain Stimulation, 10(1), 51–58.

Kronberg, G., Rahman, A., Sharma, M., Bikson, M., & Parra, L. C. (2019). Direct current stimulation boosts Hebbian plasticity in vitro. Brain Stimulation.

Kruijt, A. W., Parsons, S., & Fox, E. (2019). A meta-analysis of bias at baseline in RCTs of attention bias modification: no evidence for dot-probe bias towards threat in clinical anxiety and PTSD. Journal of Abnormal Psychology, 128(6), 563.

MacLeod, C., Mathews, A., & Tata, P. (1986). Attentional bias in emotional disorders. Journal of Abnormal Psychology, 95(1), 15.

MacLeod, C., Rutherford, E., Campbell, L., Ebsworthy, G., & Holker, L. (2002). Selective attention and emotional vulnerability: assessing the causal basis of their association through the experimental manipulation of attentional bias. Journal of Abnormal Psychology, 111(1), 107.

Martínez-Pérez, V., Castillo, A., Sánchez-Pérez, N., Vivas, A. B., Campoy, G., & Fuentes, L. J. (2019). Time course of the inhibitory tagging effect in ongoing emotional processing. A HD-tDCS study. Neuropsychologia, 135, 107242.

Mathews, A., & Mackintosh, B. (1998). A cognitive model of selective processing in anxiety. Cognitive Therapy and Research, 22(6), 539–560.

Mathews, A., & MacLeod, C. (2002). Induced processing biases have causal effects on anxiety. Cognition and Emotion, 16(3), 331–354.

Mathews, A., & MacLeod, C. (2005). Cognitive vulnerability to emotional disorders. Annual Review of Clinical Psychology, 1, 167–195.

Matsumoto, D., & Ekman, P. (1989). American-Japanese cultural differences in intensity ratings of facial expressions of emotion. Motivation and Emotion, 13(2), 143–157.

Miller, M. A., & Rahe, R. H. (1997). Life changes scaling for the 1990s. Journal of Psychosomatic Research, 43(3), 279–292.

Mogoaşe, C., David, D., & Koster, E. H. (2014). Clinical efficacy of attentional bias modification procedures: An updated meta-analysis. Journal of Clinical Psychology, 70(12), 1133–1157.

Myruski, S., Bonanno, G. A., Gulyayeva, O., Egan, L. J., & Dennis-Tiwary, T. A. (2017). Neurocognitive assessment of emotional context sensitivity. Cognitive, Affective, & Behavioral Neuroscience, 17(5), 1058–1071.

Nikolin, S., Loo, C. K., Bai, S., Dokos, S., & Martin, D. M. (2015). Focalised stimulation using high definition transcranial direct current stimulation (HD-tDCS) to investigate declarative verbal learning and memory functioning. Neuroimage, 117, 11–19.

O’Shea, J., Boudrias, M. H., Stagg, C. J., Bachtiar, V., Kischka, U., Blicher, J. U., & Johansen-Berg, H. (2014). Predicting behavioural response to TDCS in chronic motor stroke. Neuroimage, 85, 924–933.

Pergamin-Hight, L., Pine, D. S., Fox, N. A., & Bar-Haim, Y. (2016). Attention bias modification for youth with social anxiety disorder. Journal of Child Psychology and Psychiatry, 57(11), 1317–1325.

Porges, S. W. (1985). Method and apparatus for evaluating rhythmic oscillations in aperiodic physiological response systems: United States Patent Number 4,510,944.

Price, R. B., Cummings, L., Gilchrist, D., Graur, S., Banihashemi, L., Kuo, S. S., & Siegle, G. J. (2018). Towards personalized, brain-based behavioral intervention for transdiagnostic anxiety: Transient neural responses to negative images predict outcomes following a targeted computer-based intervention. Journal of Consulting and Clinical Psychology, 86(12), 1031.

Radman, T., Ramos, R. L., Brumberg, J. C., & Bikson, M. (2009). Role of cortical cell type and morphology in subthreshold and suprathreshold uniform electric field stimulation in vitro. Brain Stimulation, 2(4), 215–228.

Rotheram-Borus, M. J., Swendeman, D., & Chorpita, B. F. (2012). Disruptive innovations for designing and diffusing evidence-based interventions. American Psychologist, 67(6), 463.

Schneider, W., Eschman, A., & Zuccolotto, A. (2002). E-Prime: User’s guide. Psychology Software Incorporated.

Seibt, O., Brunoni, A. R., Huang, Y., & Bikson, M. (2015). The pursuit of DLPFC: non-neuronavigated methods to target the left dorsolateral pre-frontal cortex with symmetric bicephalic transcranial direct current stimulation (tDCS). Brain Stimulation, 8(3), 590–602.

Shahbabaie, A., Hatami, J., Farhoudian, A., Ekhtiari, H., Khatibi, A., & Nitsche, M. A. (2018). Optimizing electrode montages of transcranial direct current stimulation for attentional bias modification in early abstinent methamphetamine users. Frontiers in Pharmacology, 9.

Shaw, M. T., Kasschau, M., Dobbs, B., Pawlak, N., Pau, W., Sherman, K., … & Charvet, L. E. (2017). Remotely supervised transcranial direct current stimulation: an update on safety and tolerability. JoVE (Journal of Visualized Experiments), (128), e56211.

Shen, B., Yin, Y., Wang, J., Zhou, X., McClure, S. M., & Li, J. (2016). High-definition tDCS alters impulsivity in a baseline-dependent manner. Neuroimage, 143, 343–352.

Spielberger, C. D. (1983). Manual for the State-Trait Anxiety Inventory STAI (form Y)(“self-evaluation questionnaire”).

Sprunger, J. G. (2018). Randomized Controlled Trial of an Attention-based Intervention for Alcohol-facilitated Intimate Partner Aggression (Doctoral dissertation, Purdue University).

Tottenham, N., Tanaka, J. W., Leon, A. C., McCarry, T., Nurse, M., Hare, T. A., … & Nelson, C. (2009). The NimStim set of facial expressions: judgments from untrained research participants. Psychiatry Research, 168(3), 242–249.

Valerio, J. (2004). An FMRI study of cardiovascular reactivity to mental stress (Doctoral dissertation, Faculty of Graduate Studies, University of Western Ontario), 1–96.

Waechter, S., Stolz, J.A. (2015). Trait anxiety, state anxiety, and attentional bias to threat: Assessing the psychometric properties of response time measures. Cognitive Therapy and Research, 39, 441–458.

Wald, I., Bitton, S., Levi, O., Zusmanovich, S., Fruchter, E., Ginat, K., … & Bar-Haim, Y. (2017). Acute delivery of attention bias modification training (ABMT) moderates the association between combat exposure and posttraumatic symptoms: a feasibility study. Biological Psychology, 122, 93–97.

Weinberg, A., Venables, N. C., Proudfit, G. H., & Patrick, C. J. (2014). Heritability of the neural response to emotional pictures: evidence from ERPs in an adult twin sample. Social Cognitive and Affective Neuroscience, 10(3), 424–434

Woods, A. J., Antal, A., Bikson, M., Boggio, P. S., Brunoni, A. R., Celnik, P., … & Knotkova, H. (2016). A technical guide to tDCS, and related non-invasive brain stimulation tools. Clinical neurophysiology, 127(2), 1031–1048.

